# The QDIS-7: one scale for measuring the disease-specific quality-of-life impact of different medical conditions

**DOI:** 10.1101/2024.06.13.24308629

**Authors:** Shunichi Fukuhara, Joseph Green, Takafumi Wakita, Yosuke Yamamoto, Hajime Yamazaki, John E. Ware

## Abstract

**Background:** When studying health-related quality of life (QOL), disease-specific instruments have the advantage of measuring the unique effects of particular medical conditions. Almost every disease-specific QOL instrument uses its own metric, and measures QOL in its own content areas. The unfortunate result is that scores from different disease-specific QOL instruments cannot be compared. In contrast, the seven-item Quality of Life Disease Impact Scale (QDIS-7) has response choices on only one scale (one metric) and its content is standardized. Thus, the QDIS-7 should allow disease-specific QOL to be compared across different diseases. We therefore tested whether, unlike scores from the traditional mutually-incompatible metrics, those from the single-metric QDIS-7 are comparable across diseases.

**Methods:** Responses to the QDIS-7 questions (regarding global QOL, physical functioning, role functioning, social functioning, vitality, mental health, and health outlook) were used to compute a single score, based on an item-response model. When the QDIS-7 was completed by respondents with different diseases, the content of the question-items was the same, and the only difference was the name of the disease to which the respondents explicitly attributed any impact on their QOL. In an online survey, 2,627 adults who had sought care for headache, low-back pain, asthma, or diabetes, each responded to the QDIS-7 and to a previously-validated disease-specific QOL instrument (“legacy scale”) that was developed to measure QOL in their specific disease. We examined the slopes from four regressions of legacy-scale scores on QDIS-7 scores. Similarity of those slopes would support the hypothesis that the QDIS-7 enables quantitative comparisons of disease-specific QOL across those four different medical conditions.

**Results:** For all four groups, the regression-line slopes were nearly the same: 0.12 to 0.14 legacy-scale standard deviations per 1-point difference in QDIS-7 score. Thus, each 10-point difference in QDIS-7 scores is equal to slightly more than one standard-deviation difference in legacy-scale scores, for *all four* groups.

**Conclusions:** The relationships of score differences on the legacy measures to score differences on the QDIS-7 (i.e., the slopes) were similar across the four groups, which is consistent with the idea that the QDIS-7 enables comparisons of disease-specific QOL across different medical conditions.

## Introduction

Health-related quality of life (QOL) is generally thought of as either generic or disease-specific. Instruments for measuring generic QOL can provide information not only about patients but also about people who have not been given a diagnosis and are not undergoing medical evaluation or treatment. In contrast, an instrument for measuring disease-specific QOL provides meaningful information only about people who have the specific medical condition for which that instrument was designed. Disease-specific QOL instruments have one important advantage: They are especially sensitive and responsive to the unique effects of particular medical conditions on QOL (1) Dozens, perhaps hundreds (2, 3), of such instruments have been used in research and in daily clinical practice.

Nonetheless, disease-specific instruments have an important disadvantage: They do not facilitate comparisons across diseases. That disadvantage stems from two facts (a) they differ in the QOL domains (i.e., the content) that they measure, and (b) each instrument measures QOL on its own unique metric: The question-items (i.e., the content) and the response choices used in the Headache Impact Test differ from the question-items and the response choices used in the Asthma Control Test, etc. The result is that disease-specific QOL scores for different diseases are difficult or impossible to compare. These instruments provide no answers to questions about differences in disease-specific QOL across different medical conditions. For example, is the asthma-specific impact on QOL in people with moderately severe asthma greater or less than the diabetes-specific impact on QOL in people with moderately severe diabetes? That question cannot be answered using asthma-specific and diabetes-specific QOL instruments (4, 5), because those instruments measure QOL in different domains (content areas) and on different metrics. Similarly, in a person who has both diabetes and low-back pain, which condition has a greater impact on that person’s *disease-specific* QOL? As above, that question cannot be answered using diabetes-specific and low-back-pain-specific QOL instruments (5, 6), because those instruments measure QOL in different domains (content areas) and on different metrics.

Those considerations motivated the construction and testing of the seven-item Quality of Life Disease Impact Scale (QDIS-7) (7, 8). Unlike other disease-specific QOL instruments, the QDIS-7 uses only one set of question items, together with response choices on only one scale, to measure the QOL impact attributed by the respondent to any namable disease. Its items are sufficiently homogeneous (or unidimensional) to justify the simplicity of a 1-factor disease-specific measurement model, which consistently yields highly reliable scores quantifying QOL impact for different diseases (7, 9). It is also flexible enough to allow measurement of QOL impact attributed not only to diseases but also to symptoms, treatments, exposures, etc. Strictly speaking, rather than being merely disease-specific, the QDIS-7 is attribution-specific.

The QDIS-7 is *sui generis* in two ways. To the best of our knowledge, no other instrument allows comparisons of disease-specific QOL impacts between individuals with different health conditions, and no other instrument allows comparisons of disease-specific QOL impacts of different health conditions within a single individual.

Given the potential advantages of the QDIS-7 over the many disease-specific measures of QOL that are already in widespread use (i.e., “legacy” measures), and also given the increasing prevalence and importance of multimorbidity in rapidly aging populations, this new instrument has the potential to be useful worldwide for multiple purposes. Proliferation of the QDIS-7 will require further evidence that it can be used to quantify the disease-specific magnitudes of QOL impact across diseases in meaningfully comparable standardized units of QOL, in various social-cultural contexts.

With that in mind, we compared the QDIS-7 with legacy measures of disease-specific QOL in people with headache, low-back pain, asthma, and diabetes, and we tested whether the disease-specific QOL impacts of those four different medical conditions could be quantified on a single metric.

## Methods

### Participants, and minimization of potential bias

For this cross-sectional-study, the participants were in four online panels. Because these QOL data come from self-reports, bias might have been introduced if the participants’ identities were known to the researchers, and particularly if their identities were known to people involved in their health care. Those possibilities were minimized by having a third-party survey-research company (Cross Marketing Inc.) assemble the online panels, collect the data, and anonymize the data to ensure that the researchers did not know the participants’ identities, and that the participants identities were also not known to anyone involved in their health care. The anonymization procedures were described in the informed-consent document that was distributed to all participants. The survey-research company also ensured that in this online survey there were no missing data.

The research was planned to include women and men aged 16-84 years of age. Balancing budgetary constraints against the need to acquire enough data for valid and reliable comparisons and psychometric tests, each of the four online panels had at least 500 participants. Each panel comprised people who reported having sought care for one of four medical conditions: headache, low-back pain, asthma, or diabetes. Basic demographic information is in Table 1.

**Table 1.**
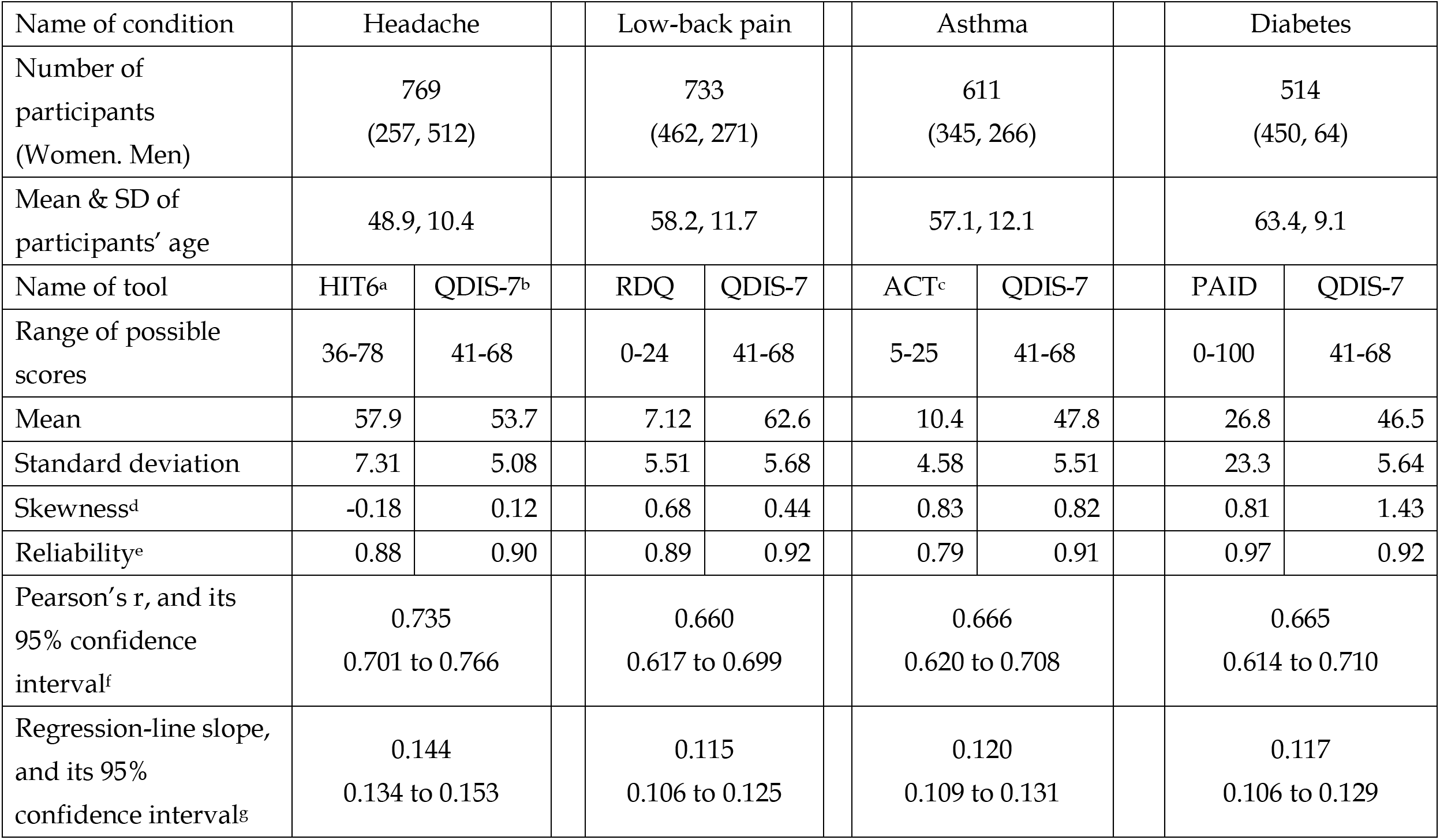
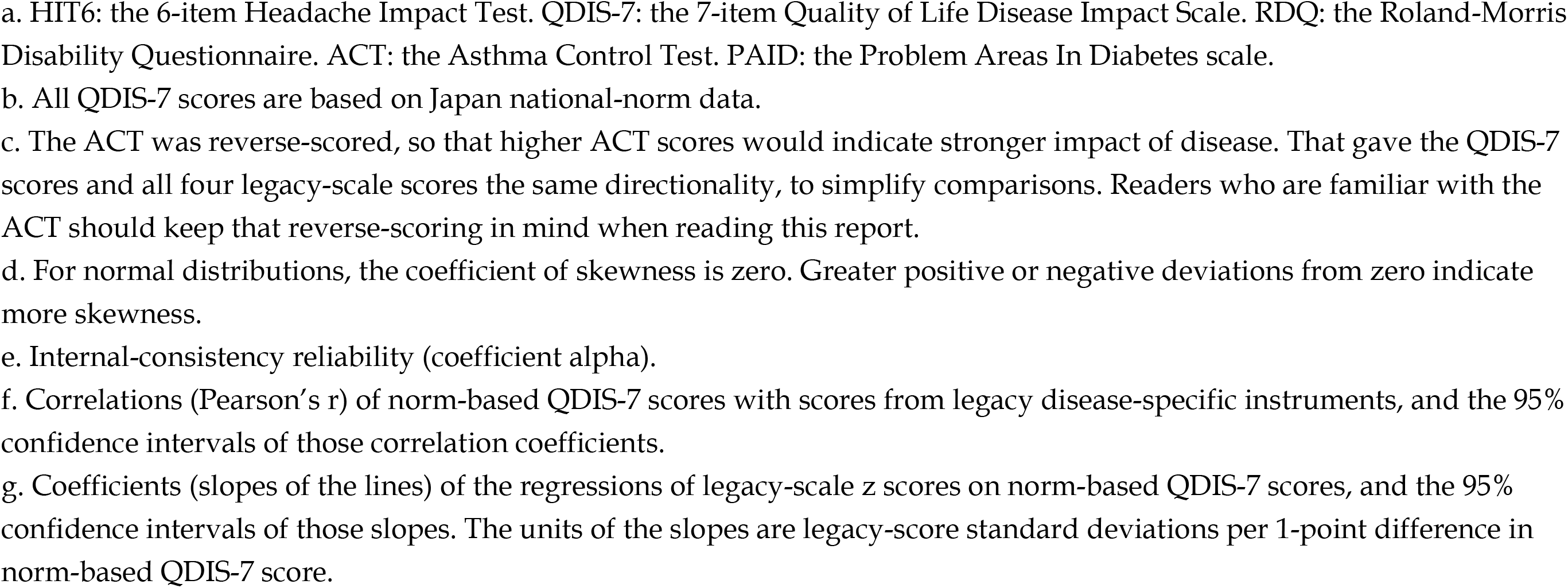
Summary information on participants and disease-specific quality-of-life impact measures of four different medical conditions.

| Name of condition | Headache |  | Low-back pain |  | Asthma |  | Diabetes |  |
| --- | --- | --- | --- | --- | --- | --- | --- | --- |
| Number of participants<br>(Women. Men) | 769<br>(257, 512) |  | 733<br>(462, 271) |  | 611<br>(345, 266) |  | 514<br>(450, 64) |  |
| Mean & SD of participants' age | 48.9, 10.4 |  | 58.2, 11.7 |  | 57.1, 12.1 |  | 63.4, 9.1 |  |
| Name of tool | HIT6<sup>a</sup> | QDIS-7<sup>b</sup> | RDQ | QDIS-7 | ACT<sup>c</sup> | QDIS-7 | PAID | QDIS-7 |
| Range of possible scores | 36-78 | 41-68 | 0-24 | 41-68 | 5-25 | 41-68 | 0-100 | 41-68 |
| Mean | 57.9 | 53.7 | 7.12 | 62.6 | 10.4 | 47.8 | 26.8 | 46.5 |
| Standard deviation | 7.31 | 5.08 | 5.51 | 5.68 | 4.58 | 5.51 | 23.3 | 5.64 |
| Skewness<sup>d</sup> | -0.18 | 0.12 | 0.68 | 0.44 | 0.83 | 0.82 | 0.81 | 1.43 |
| Reliability<sup>e</sup> | 0.88 | 0.90 | 0.89 | 0.92 | 0.79 | 0.91 | 0.97 | 0.92 |
| Pearson's r, and its 95% confidence interval<sup>f</sup> | 0.735<br>0.701 to 0.766 |  | 0.660<br>0.617 to 0.699 |  | 0.666<br>0.620 to 0.708 |  | 0.665<br>0.614 to 0.710 |  |
| Regression-line slope, and its 95% confidence intervals<sup>g</sup> | 0.144<br>0.134 to 0.153 |  | 0.115<br>0.106 to 0.125 |  | 0.120<br>0.109 to 0.131 |  | 0.117<br>0.106 to 0.129 |  |

- a. HIT6: the 6-item Headache Impact Test. QDIS-7: the 7-item Quality of Life Disease Impact Scale. RDQ: the Roland-Morris Disability Questionnaire. ACT: the Asthma Control Test. PAID: the Problem Areas In Diabetes scale. - b. All QDIS-7 scores are based on Japan national-norm data. - c. The ACT was reverse-scored, so that higher ACT scores would indicate stronger impact of disease. That gave the QDIS-7 scores and all four legacy-scale scores the same directionality, to simplify comparisons. Readers who are familiar with the ACT should keep that reverse-scoring in mind when reading this report. - d. For normal distributions, the coefficient of skewness is zero. Greater positive or negative deviations from zero indicate more skewness. - e. Internal-consistency reliability (coefficient alpha). - f. Correlations (Pearson's $r$ ) of norm-based QDIS-7 scores with scores from legacy disease-specific instruments, and the 95% confidence intervals of those correlation coefficients. - g. Coefficients (slopes of the lines) of the regressions of legacy-scale $z$ scores on norm-based QDIS-7 scores, and the 95% confidence intervals of those slopes. The units of the slopes are legacy-score standard deviations per 1-point difference in norm-based QDIS-7 score.

### The QDIS-7

The QDIS-7 is uniquely flexible and adaptable because of the structure of its question-items and the uniformity of its response choices (7, 8, 10). Briefly, each question-item refers to one aspect of the respondents’ QOL, and it asks the respondents to rate the QOL impact that they attribute to a named condition. To do that, each question-item uses a fill-in-the-attribution (fill-in-the-condition) structure. For example, one of the QDIS-7 question-items is “In the past 4 weeks, how often did [CONDITION] limit your usual social activities with family, friends, or others close to you?”. For participants who have headache, [CONDITION] is replaced with “headache”. For those who have diabetes, [CONDITION] is replaced with “diabetes”, etc. (7). Translation and psychometric testing of the QDIS-7 are described in Appendix 1.

### Disease-specific QOL instruments for each of the medical conditions studied

After responding to the QDIS-7, each participant also responded to a well-established (i.e., “legacy”) disease-specific instrument for the medical condition that they reported having: Those with headache responded to the 6-item Headache Impact Test (HIT6 (11, 12)), those with low-back pain responded to the 24-item Roland-Morris Disability Questionnaire (RDQ (6, 13)), those with asthma responded to the 5-item Asthma Control Test (ACT (4, 14)), and those with diabetes responded to the 20-item Problem Areas In Diabetes scale (PAID (5, 15)).

### Analyses

The QDIS-7 scores reported here are norm-based. Norm-based scoring began with a sample of the population of Japan. That sample was recruited and studied separately from the present study’s participants described above. The methods used to derive the norm-based scores are described in Appendix 2.

For each of the four groups, we computed the norm-based QDIS-7 scores and the scores on the applicable legacy disease-specific measure (i.e., HIT6, RDQ, ACT, or PAID). For the legacy disease-specific measures, scores were computed as previously reported for each instrument (4–6, 11). For all further computations, we reverse-scored the ACT, so that higher ACT scores would indicate stronger impact of disease. That gave the QDIS-7 scores and all four legacy scores the same directionality, which simplifies comparisons. Readers who are familiar with the ACT should keep that reverse-scoring in mind when reading this report.

We computed Pearson’s r for each of the four groups, to quantify the strength of the associations between legacy-scale scores and QDIS-7 scores. We also computed internal-consistency reliability (alpha) and examined the frequency distributions of legacy-scale and QDIS-7 scores..

One important question is whether the QDIS-7 indeed allows the disease-specific QOL impact of different medical conditions to be quantified on the same scale. To answer that question, we hypothesized that the differences in legacy scores associated with a unit difference in QDIS-7 scores are consistent across the four groups. Testing that hypothesis required computing the slope of the relationship between legacy scores and QDIS-7 scores for each group, and then comparing those slopes among the four groups. First we standardized the legacy scores within each of the four groups, such that all four distributions of legacy scores had the same mean (i.e., zero) and the same standard deviation (i.e., 1). The legacy scores thus became z scores. Linear regression of those z scores on the norm-based QDIS-7 scores provided the slopes used to test the hypothesis posed above.

### Patient and public involvement

Neither patients nor the general public were involved in planning or carrying out this study.

## Results

### Participants

A total of 2,627 people responded to the QDIS-7 and to one of the legacy instruments. Table 1 shows the numbers of participants in each of the four groups, along with basic demographic information.

### Descriptive statistics

The means of legacy QOL measures differ but cannot be meaningfully compared, but all of the QDIS-7 means are on the same metric, so they can be compared. They show that the QOL impacts of the four conditions were, from highest to lowest, low-back pain > headache > asthma > diabetes. The QOL impact of low-back pain was 1.61 standard deviations greater than that of diabetes.

**Correlations, reliability, and frequency distributions (Figure 1**, **Table 1) Correlations:** The correlations (Pearson’s r) between QDIS-7 scores and legacy scale scores ranged from 0.660 to 0.735.

**Figure 1.**
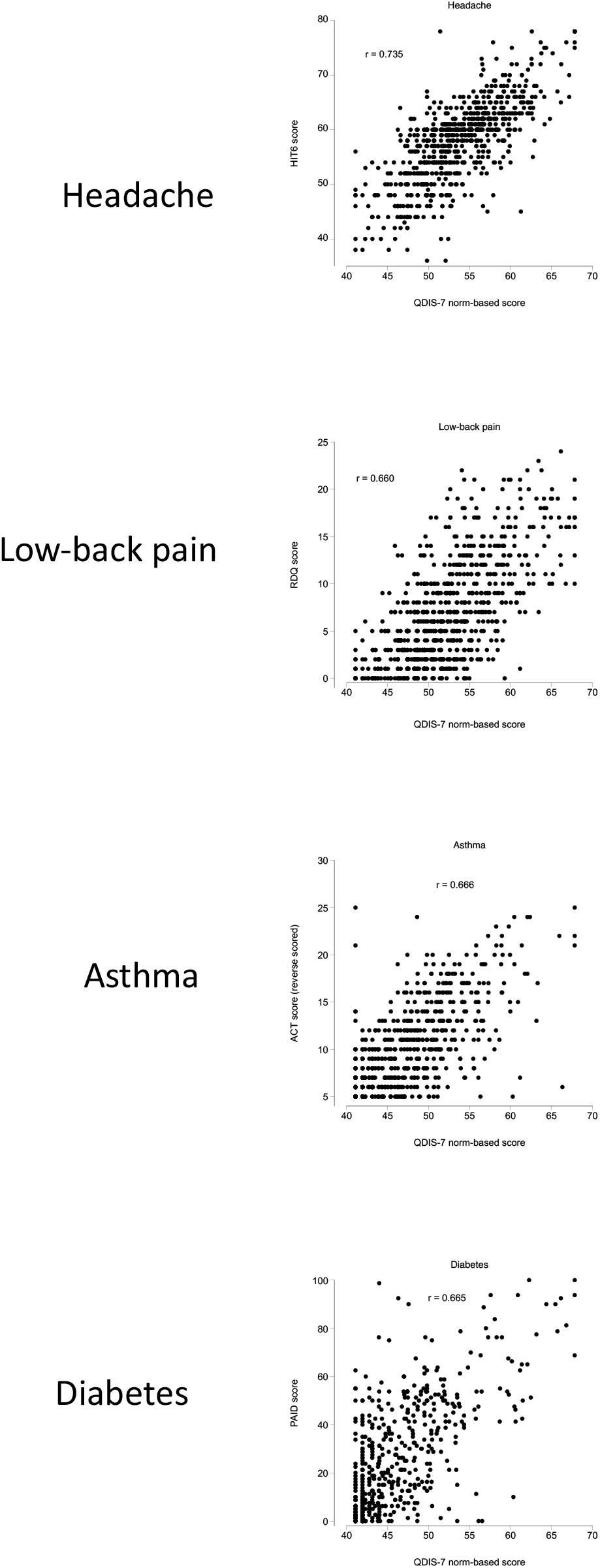
Scatterplots showing the relationships between the norm-based scores on the QDIS-7 and the scores on the four legacy disease-specific scales.

**Reliability:** In all four groups, the alpha of the QDIS-7 scores was at least 0.90.

**Headache:** In the group with headache, the frequency distributions of both HIT6 scores and QDIS-7 scores were approximately normal.

**Low-back pain:** The distribution of RDQ scores was right-skewed. In contrast, the distribution of QDIS-7 scores was closer to normal.

**Asthma:** The ACT scores were very strongly left-skewed (after reverse-scoring), while the QDIS-7 scores were slightly less skewed.

**Diabetes:** Both the PAID scores and the QDIS-7 scores were markedly right-skewed.

### Consistency of the magnitude of relationships between QDIS-7 scores and legacy-scale scores across medical conditions

Figure 3 shows the results of linear regression of legacy-scale z scores on norm-based QDIS-7 scores, for the four groups. The slopes of the four regression lines varied over a small range (Table 1): from 0.115 to 0.144 legacy scale standard deviations per 1-point difference in QDIS-7 norm-based score, for all four groups.

## Discussion

### Summary of main findings

Despite the differences in content (i.e., differences in domain coverage) between the QDIS-7 and the legacy scales studied, the correlations between their scores were substantial (Table 1), which is consistent with the presence of a large common underlying disease-specific QOL-impact factor. All of the QDIS-7 scores were highly reliable. For three of the four medical conditions studied, the frequency distributions of QDIS-7 scores were less skewed than those of the legacy-scale scores (Figure 2, Table 1). Thus, using the QDIS-7 should simplify both the interpretation of group-level statistics (mean, standard deviation, etc.) and the interpretation of individual scores in relation to those group means, standard deviations, etc. Figure 3 shows a particularly important finding: The relation between QDIS-7 scores and legacy-scale scores (i.e., the slope) was consistent across the four medical conditions studied.

**Figure 2.**
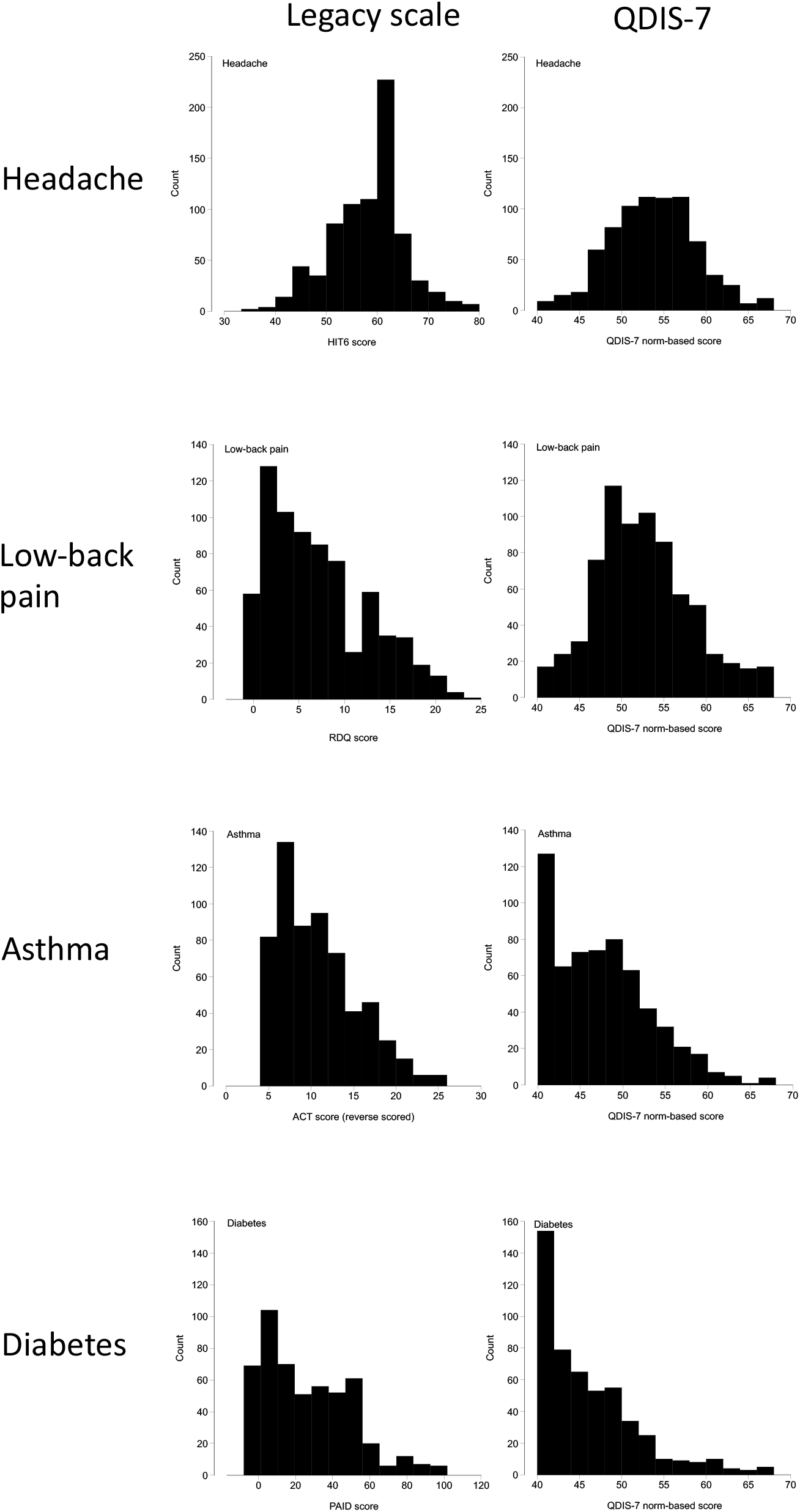
Frequency distributions of the norm-based scores on the QDIS-7 and the scores on the four legacy disease-specific scales.

### Advantages over generic instruments

Using generic health-related QOL tools instead of disease-specific tools entails a loss of information. For example, if a person has asthma, and that person’s generic health-related QOL is low, we do not know why it is low. Specifically, we do not know whether it is low because of their asthma or because of something else related to their health. With generic health-related QOL tools, QOL impacts are attributed to physical or mental health in general, not to a particular, named condition. In contrast, information about QOL impacts attributed to a particular condition is provided by instruments for measuring disease-specific QOL, of which the QDIS-7 is one. Furthermore, in people with multimorbidity, generic health-related QOL tools cannot provide information about the relative QOL impacts of each of the various comorbid conditions. As noted above, such information can be acquired only by measuring attribution-specific QOL, such as with the QDIS-7 (9).

### Consistency of the relation between QDIS-7 scores and legacy-scale scores across diseases (Figure 3)

We also tested the hypothesis that the differences in legacy scores that are associated with a unit difference in QDIS-7 scores are consistent across the four groups. The results shown in Figure 3 support that hypothesis. In all four groups, each 10-point difference in QDIS-7 score was associated with a 1.2-to-1.4-SD difference in the score on the legacy instrument. That is, the range over which the regression-line slopes varied was remarkably small. That similarity among the four slopes has an important implication. It supports the use of a single scale, the QDIS-7 in norm-score units, instead of the different and mutually-incompatible legacy scales in their original units.

**Figure 3.**
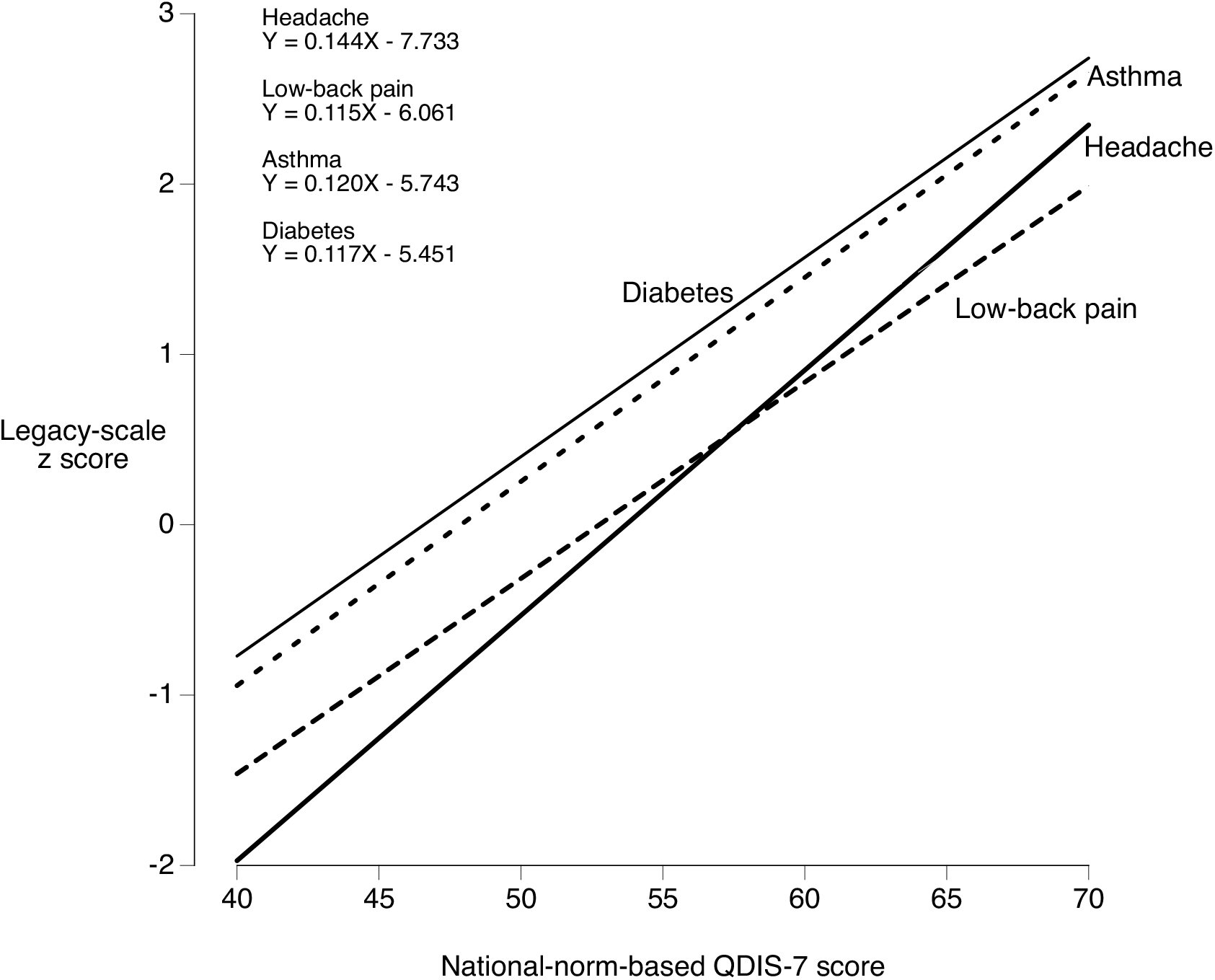
Results of regression of legacy-scale z scores on norm-based QDIS-7 scores. The slopes of the regression lines varied from 0.115 to 0.144 legacy score standard deviations per 1-point difference in norm-based QDIS-7 score. Confidence intervals of the slopes are in Table 1.

This might appear to be similar to the standardized mean difference, which puts different QOL measures on a single scale (16). However, as described in Appendix 3, that similarity is superficial.

Using the QDIS-7 instead of mutually-incompatible legacy scales also has an advantage when comparing scores longitudinally. For example, with QDIS-7 scores, a 10-point improvement caused by treatment of headache would be on the same scale as a 10-point improvement caused by treatment of asthma. Of course, whether those two 10-point improvements would justify two similar clinical decisions is a separate question. Relations between QOL and clinical decisions can be addressed with different methods, such as by computing minimally important changes (MIC) for each of the diagnoses in question (17, 18), which is beyond the scope of this study.

### Using the QDIS-7 to answer new questions

Among disease-specific QOL instruments, the QDIS-7 is uniquely useful. It allows clinicians, researchers, policy makers, etc., to answer important questions by making comparisons that were not possible previously. For example, consider a person who has both diabetes and asthma: Of those two conditions, which has more impact on that person’s QOL? Neither generic QOL instruments nor legacy disease-specific QOL instruments can answer that question, but the QDIS-7 can. Similarly, suppose health-policy researchers compare a group of people with headache to a group with low-back pain. If those researchers are interested only in generic QOL, then they can simply use, for example, the SF-36. But what if they are interested in disease-specific QOL? That is, what if they ask: In which of those two groups is the disease-specific impact on QOL greater? Again, such a question cannot be answered either by using generic QOL instruments or by using legacy disease-specific QOL instruments, but it can be answered by using the QDIS-7. The results of the present study offer a concrete example: *Which of the four groups had the highest disease-specific QOL and which had the lowest? And how large was the difference in disease-specific QOL between those two groups?* As noted above, the mean QDIS-7 scores (Table 1) show that the disease-specific impact on QOL was highest for those with low-back pain and it was lowest for those with diabetes: The mean QDIS-7 scores in those two groups were 62.6 and 46.5, respectively. (This is consistent with the fact that all people with low-back pain are symptomatic by definition, whereas some people with diabetes are asymptomatic.) That difference is 16.1 points on the QDIS-7 scale. With norm-based scoring, a difference of 10 points is 1 standard deviation, so a difference of 16.1 points is 1.61 standard deviations. Differences of 1.5 standard deviations are considered to be very large (19, 20). This answers the two questions posed above (in *italics*), and it illustrates how the QDIS-7 can be used to answer questions about disease-specific QOL that previously were difficult or impossible to address.

### The QDIS-7 is flexible and adaptable

Unlike generic QOL instruments, the QDIS-7 was constructed to be used primarily with data from people who have a known diagnosis. In that sense, it is similar to legacy disease-specific instruments. Going beyond that similarity, the QDIS-7 can be used to measure not only the QOL effects of any specifiable disease, but also those of almost any other namable status or condition to which an impact on QOL might be attributed. Its unique fill-in-the-attribution structure should make the QDIS-7 more flexible and adaptable than any other QOL instrument of which we are aware. Specifically, in addition to measuring the impact of diseases, the QDIS-7 can also measure the impact of symptoms (headache and low-back pain in the present study), treatments (21, 22), exposures (23, 24), and any other specifiable (25) status or condition that might affect regular daily physical, psychological, or social functioning. Applications of the QDIS-7 may include QOL associated with environmental pollution, healthcare workers’ QOL, maternal postpartum QOL, and caregivers’ QOL, (23, 24, 26–34). None of those examples is a disease, but the QDIS-7 can be used to quantify the specific impact of each one on QOL, using the same metric as for asthma, diabetes, etc.

Furthermore, *de novo* development and testing of patient-reported outcome measures can be very time-consuming and resource-intensive. In the presence of new and emerging medical conditions, new disease-specific QOL instruments suddenly become needed, as occurred during the COVID-19 pandemic (26, 35). In such situations, the QDIS-7 could be adapted to meet those new needs quickly and easily.

### Summary of the main advantage of the QDIS-7

Both the QDIS-7 and legacy disease-specific QOL measures have the advantage of being particularly sensitive to disease-specific impacts on QOL. For legacy measures, that advantage comes with an important limitation: Impacts on QOL cannot be compared across diseases. The QDIS-7 has no such limitation. The QDIS-7’s unique advantage is that it allows quantitative comparisons of attribution-specific impacts on QOL across diseases, exposures, symptoms, treatments, etc.

### Limitations, and directions for further research

First, we used only one legacy disease-specific measure for each of the four disease groups studied, but some diseases have many legacy disease-specific QOL instruments, which might vary in their relationship to the QDIS-7.

Second, we studied only four medical conditions. Those four were chosen in anticipation of using them in future work comparing QDIS-7 scores across countries, and because they are commonly encountered in primary care settings, and also because they are relatively likely to affect QOL (unlike, for example, hypertension). Useful insights could certainly come from comparing QDIS-7 scores in a larger group of medical conditions.

Third, as noted above, the QDIS-7 is uniquely appropriate for standardizing attribution-specific QOL in people with more than one medical condition. More work is needed on the usefulness of the QDIS-7 in people with multimorbitiies. In that regard, recent cross-sectional (9) and longitudinal (25) studies have shown that people with multimorbitiies can distinguish between the impacts of each of their conditions on QOL.

Fourth, although this study was cross-sectional, disease-specific instruments are particularly responsive to changes over time (10, 36, 37). The comparative responsiveness of the QDIS-7, legacy instruments, and generic instruments is a topic for future study.

Fifth, as noted above, answering questions regarding clinical interpretation and clinical decisions requires data on the MICs of QDIS-7 scores (17, 18).

## Conclusion

For measuring disease-specific QOL (in fact, attribution-specific QOL), the QDIS-7 has an important advantage over legacy tools. It is the only QOL measure we know of that uses a single metric to quantify the QOL impact attributed to specific diseases, symptoms, treatments, exposures, etc. It enables, for the first time, quantitative comparisons across different attributions. Using the QDIS-7, researchers and clinicians can now answer questions that previously were difficult or impossible to address.

## Data Availability

All reasonable requests for access to the data produced in the present study should be directed to the corresponding author.

## Ethics review, funding, reporting, and availability

On August 21, 2020, the plan for this study (201611-3) was approved by the Institutional Review Board of the the Institute for Health Outcomes and Process Evaluation Research (iHope: <http://www.i-hope.jp/>). Funding for parts of this study came from the Ministry of Education, Culture, Sports, Science and Technology of Japan, through the Japan Society for the Promotion of Science (18H03024). We used the STROBE checklist for cross-sectional studies when writing this report (38). The QDIS-7 is available after registration through. It is available royalty-free for academic users.

## Appendix 1

### Translation of the QDIS-7 into Japanese, and initial psychometric testing

The Japanese-language version of the QDIS-7 resulted from the commonly-used procedure of multiple forward translations, reconciliation, back-translation, and consultation with the developer of the original (English-language) version. The translators involved in the forward translations were native speakers of Japanese. Working independently, their goal was to produce a version of the QDIS-7 that would use colloquial language and would be easily understood by adults whose native language was Japanese.

All but one of the response choices in the original (English-language) version had been used in other QOL instruments for which Japanese-language versions already existed. In those cases, the Japanese-language version of the QDIS-7 included Japanese-language response choices that were already being used successfully in other QOL instruments.

The translator involved in the back-translation was a native speaker of English. The back-translated version was used in consultations with the developer of the original (English-language) version.

Initial psychometric evaluation of the QDIS-7 involved factor analysis, computation of internal-consistency reliability, and criterion-related validation testing. Quantitative details have been reported (21), and the overall findings are summarized here. The results of factor analysis confirmed the QDIS-7’s hypothesized unidimensionality: More than half of the variance was explained by the first factor, and all of the factor loadings were high. Internal-consistency reliability of QDIS-7 scores was also high (21), as it was in the present study (Table 1).

Criterion-based validation testing showed that, as hypothesized for a disease-specific QOL instrument, in patients who had chronic kidney disease (CKD) the sensitively of the QDIS-7 to CKD stage was greater than that of a generic QOL instrument (21).

## Appendix 2

### Norm-based scoring

Norm-based scoring began with data from 3,131 people in an age-and-gender-stratified representative sample of the population of Japan.

It is important to note that these were not the people in the four groups in the main study. That is, they were not the people who provided the data used to make Table 1 and Figures 1, 2, and 3.

Among those 3,131 people, 2,659 reported having at least one of the 41 medical conditions listed below. The QDIS-7 scores of those 2,659 people were used as the basis for the norm-based scoring.

1. Hypertension
2. Myocardial infarction experienced in the past year
3. Congestive heart failure or cardiomegaly
4. Diabetes or high blood sugar
5. Angina pectoris
6. Stroke or other cerebrovascular disorders
7. Cancer (excluding skin cancer)
8. Asthma
9. Hypothyroidism or diseases causing thyroid dysfunction
10. Chronic allergy or sinusitis
11. Atopic dermatitis (eczema)
12. Chronic skin diseases other than atopic dermatitis
13. Chronic obstructive pulmonary disease (COPD)
14. Kidney disease
15. Rheumatoid arthritis
16. Connective tissue diseases (systemic lupus erythematosus, scleroderma, etc.)
17. Osteoarthritis or degenerative joint disease
18. Osteoporosis
19. Stomach ulcers, gastritis, duodenitis, or other gastric diseases
20. Hepatitis B or hepatitis C
21. Irritable bowel syndrome
22. Inflammatory bowel disease (ulcerative colitis, Crohn’s disease)
23. Obesity
24. Anemia
25. Depression
26. Chronic fatigue syndrome
27. Migraine
28. Headaches other than migraine
29. Prostate enlargement
30. Erectile dysfunction (ED)
31. Gynecological diseases (uterus, ovaries, etc.)
32. Seasonal allergies such as hay fever
33. Chronic back pain or sciatica
34. Difficulty or inability to see even with glasses or contact lenses
35. Difficulty or inability to hear in one or both ears
36. Problems with upper or lower limbs (amputation, paralysis, weakness, etc.)
37. Joint problems in the ankle or toes
38. Joint problems in the hip or knee
39. Urinary problems such as urinary incontinence or difficulty urinating
40. Shoulder pain or inflammation
41. Coldness in the back or legs

Participants who reported having more than one of those 41 medical conditions were asked which one had the greatest effect on their QOL. Then they responded to the QDIS-7 with regard to that one medical condition.

For example, suppose a participant reported having both asthma and diabetes, and also reported that, of those two medical conditions, asthma had the greater effect on their QOL. In that case, the participant would respond to the QDIS-7 items in asthma-specific form.

The result was a set of QDIS-7 data with attributions to various medical conditions, from 2,659 people.

Using those data and a 2-parameter partial-credit model based on item-response theory, we computed the mean θ of each of the five response categories, for each of the QDIS-7 question items.

Because of the nature of the item-response model, for each QDIS-7 item the mean of the five responses’ locations was 0 and the standard deviation was 1. Those were transformed to 50 and 10, respectively, and then the mean of the seven QDIS-7 items was the norm-based QDIS-7 score. Thus, each 1-point difference in norm-based QDIS-7 scores is a difference of 0.1 standard deviations.

## Appendix 3

### Differences between norm-based QDIS-7 scores and the standardized mean difference

The QDIS-7 might appear to be similar to the standardized mean difference (SMD) (15), because both Figure 3 and the SMD put different QOL measures on a single scale, but that similarity is superficial. The SMD is commonly used in meta-analyses, to combine results from different studies that “all assess the same outcome, but measure it in a variety of ways” (16). Its denominator is the standard deviation of a specified outcome among the participants in the studies being meta-analyzed. In contrast, norm-based QDIS-7 scores are derived not from a group with a particular disease, but from the chronically-ill population as a whole (Appendix 2). The QDIS-7 quantifies QOL impact in attribution-specific population-based units, rather than in units based on a particular set of previously-published studies. Furthermore, using the SMD in meta-analyses assumes that the difference being quantified is between two measurements of the same domain. In contrast, the QDIS-7 is unidimensional (7, 21), so issues regarding domains do not arise.

## References

1. Patrick DL, Deyo RA. Generic and disease-specific measures in assessing health status and quality of life. Med Care. 1989 Mar;27(3 Suppl):S217–32. doi: 10.1097/00005650-198903001-00018. PMID: 2646490.

2. Fayers PM, Machin D. Quality of life: the assessment, analysis and reporting of patient-reported outcomes: John Wiley & Sons; 2015.

3. Nevarez-Flores AG, Chappell KJ, Morgan VA, Neil AL. Health-Related Quality of Life Scores and Values as Predictors of Mortality: A Scoping Review. J Gen Intern Med. 2023;38(15):3389–405.

4. Nathan RA, Sorkness CA, Kosinski M, Schatz M, Li JT, Marcus P, et al. Development of the asthma control test: a survey for assessing asthma control. J Allergy Clin Immunol. 2004;113(1):59–65.

5. Polonsky WH, Anderson BJ, Lohrer PA, Welch G, Jacobson AM, Aponte JE, et al. Assessment of diabetes-related distress. Diabetes care. 1995;18(6):754–60.

6. Roland M, Morris R. A study of the natural history of back pain. Part I: development of a reliable and sensitive measure of disability in low-back pain. Spine (Phila Pa 1976). 1983;8(2):141–4.

7. Ware JE, Jr., Gandek B, Guyer R, Deng N. Standardizing disease-specific quality of life measures across multiple chronic conditions: development and initial evaluation of the QOL Disease Impact Scale (QDIS**^®^**). Health Qual Life Outcomes. 2016;14:84.

8. Ware JE, Jr., Gandek B, Allison J. The Validity of Disease-specific Quality of Life Attributions Among Adults with Multiple Chronic Conditions. Int J Stat Med Res. 2016;5(1):17–40.

9. Ware JE. Factor analysis supports the validity of disease-specific health-related quality of life (QOL) measures among US population adults with comorbid conditions. Quality of Life Research. 2023;32:S106.

10. Ware JE, Jr., Richardson MM, Meyer KB, Gandek B. Improving CKD-Specific Patient-Reported Measures of Health-Related Quality of Life. J Am Soc Nephrol. 2019;30(4):664–77.

11. Kosinski M, Bayliss MS, Bjorner JB, Ware JE, Jr., Garber WH, Batenhorst A, et al. A six-item short-form survey for measuring headache impact: the HIT-6. Qual Life Res. 2003;12(8):963–74.

12. Ohbu S, Igarashi H, Okayasu H, Sakai F, Green J, Heller RF, et al. Development and testing of the Japanese version of the migraine-specific quality of life instrument. Qual Life Res. 2004;13(8):1489–93.

13. Suzukamo Y, Fukuhara S, Kikuchi S, Konno S, Roland M, Iwamoto Y, et al. Validation of the Japanese version of the Roland-Morris Disability Questionnaire. Journal of orthopaedic science : official journal of the Japanese Orthopaedic Association. 2003;8(4):543–8.

14. Ueno S, Obase Y, Ohfuji T, Shimizu H, Sugiu T, Ohue Y, et al. [Asthma control test (ACT) faultiness and the remediation by peak flow (PEF) measurement at outpatient department]. Arerugi. 2008;57(7):862–71.

15. Ishii H. The Japanese version of problem area in diabetes scale : a clinical and research tool for the assessment of emotional functioning among diabetic patients (abstract). Diabetes (New York, NY). 1999;48(1).

16. Higgins JPT, Li T, Deeks JJ (editors). Chapter 6: Choosing effect measures and computing estimates of effect. In: Higgins JPT, Thomas J, Chandler J, Cumpston M, Li T, Page MJ, Welch VA (editors). Cochrane Handbook for Systematic Reviews of Interventions version 6.4 (updated August 2023). Cochrane, 2023. Available from www.training.cochrane.org/handbook and https://training.cochrane.org/handbook/current/chapter-06. Accessed on January 25, 2024.

17. Tsujimoto Y, Fujii T, Tsutsumi Y, Kataoka Y, Tajika A, Okada Y, Carrasco-Labra A, Devji T, Wang Y, Guyatt GH, Furukawa TA. Minimal important changes in standard deviation units are highly variable and no universally applicable value can be determined. J Clin Epidemiol. 2022 May;145:92–100. doi: 10.1016/j.jclinepi.2022.01.017. Epub 2022 Jan 25. PMID: 35091045.

18. Wyrwich KW, Norman GR. The challenges inherent with anchor-based approaches to the interpretation of important change in clinical outcome assessments. Qual Life Res. 2023 May;32(5):1239–1246. doi: 10.1007/s11136-022-03297-7. Epub 2022 Nov 18. PMID: 36396874.

19. Cohen J. (1988). Statistical Power Analysis for the Behavioral Sciences. New York, NY: Routledge Academic

20. Sullivan GM, Feinn R. Using Effect Size-or Why the P Value Is Not Enough. J Grad Med Educ. 2012 Sep;4(3):279–82. doi: 10.4300/JGME-D-12-00156.1. PMID: 23997866; PMCID: PMC3444174.

21. Fukuhara S, Yamazaki H, Wakita T, Ware JJE, Wang J, Onishi Y, et al. Validation of a new instrument for measuring disease-specific quality of life: A pilot study among patients with chronic kidney disease and hyperkalemia. Annals of Clinical Epidemiology, 2023, Volume 5, Issue 1, Pages 13–19. 10.37737/ace.23003

22. Shibagaki Y, Yamazaki H, Wakita T, Ware JE, Wang J, Onishi Y, et al. Impact of treatment of hyperkalaemia on quality of life: design of a prospective observational cohort study of long-term management of hyperkalaemia in patients with chronic kidney disease or chronic heart failure in Japan. BMJ Open. 2023;13(12):e074090.

23. Yamazaki S, Nitta H, Murakami Y, Fukuhara S. Association between ambient air pollution and health-related quality of life in Japan: ecological study. International journal of environmental health research. 2005;15(5):383–91.

24. Singh G, Prakash J, Ray SK, Yawar M, Habib G. Development and evaluation of air pollution-linked quality of life (AP-QOL) questionnaire: insight from two different cohorts. Environ Sci Pollut Res Int. 2021 Aug;28(32):43459–43475. doi: 10.1007/s11356-021-13754-4. Epub 2021 Apr 9. PMID: 33835344.

25. McEntee ML, Gandek B, Ware JE. Improving multimorbidity measurement using individualized disease-specific quality of life impact assessments: predictive validity of a new comorbidity index. Health Qual Life Outcomes. 2022;20(1):108.

26. Yamamoto R, Yamazaki H, Kobara S, Iizuka H, Hijikata Y, Miyashita J, et al. Development and Initial Psychometric Validation of the COVID-19 Pandemic Burden Index for Healthcare Workers. J Gen Intern Med. 2023;38(5):1239–47.

27. Van Laar D, Edwards JA, Easton S. The Work-Related Quality of Life scale for healthcare workers. J Adv Nurs. 2007 Nov;60(3):325–33. doi: 10.1111/j.1365-2648.2007.04409.x. PMID: 17908128.

28. Mishina H, Hayashino Y, Fukuhara S. Test performance of two-question screening for postpartum depressive symptoms. Pediatrics international : official journal of the Japan Pediatric Society. 2009;51(1):48–53.

29. Hill PD, Aldag JC, Hekel B, Riner G, Bloomfield P. Maternal Postpartum Quality of Life Questionnaire. J Nurs Meas. 2006 Winter;14(3):205–20. doi: 10.1891/jnm-v14i3a005. PMID: 17278340.

30. Mokhtaryan-Gilani, T., Kariman, N., Nia, H.S. et al. The Maternal Postpartum Quality of Life Instrument (MPQOL-I): development and psychometric evaluation in an exploratory sequential mixed-method study. BMC Pregnancy Childbirth 22, 576 (2022). 10.1186/s12884-022-04900-y

31. Miyashita M, Yamaguchi A, Kayama M, Narita Y, Kawada N, Akiyama M, et al. Validation of the Burden Index of Caregivers (BIC), a multidimensional short care burden scale from Japan. Health Qual Life Outcomes. 2006;4:52.

32. Novak M, Guest C. Application of a multidimensional caregiver burden inventory. Gerontologist. 1989 Dec;29(6):798–803. doi: 10.1093/geront/29.6.798. PMID: 2516000.

33. Joseph, S., Becker, S., Elwick, H., & Silburn, R. (2012). Adult carers quality of life questionnaire (AC-QoL): Development of an evidence-based tool. Mental Health Review Journal, 17(2), 57–69. 10.1108/13619321211270380

34. Martin MP, McEntee ML, Suri Y. Caregiver Quality of Life: How to Measure It and Why. American Journal of Health Promotion. 2021;35(7):1042–1045. doi:10.1177/08901171211030142f

35. Amdal, C.D., Taylor, K., Kuliś, D. et al. Health-related quality of life in patients with COVID-19; international development of a patient-reported outcome measure. J Patient Rep Outcomes 6, 26 (2022).

36. Wiebe S, Guyatt G, Weaver B, Matijevic S, Sidwell C. Comparative responsiveness of generic and specific quality-of-life instruments. J Clin Epidemiol. 2003 Jan;56(1):52–60. doi: 10.1016/s0895-4356(02)00537-1. PMID: 12589870.

37. Puhan MA, Guyatt GH, Goldstein R, Mador J, McKim D, Stahl E, Griffith L, Schunemann HJ: Relative responsiveness of the Chronic Respiratory Questionnaire, St. Georges Respiratory Questionnaire and four other health-related quality of life instruments for patients with chronic lung disease. Respiratory medicine 2007, 101(2):308–316. 10.1016/j.rmed.2006.04.023

38. Elm E v, Altman D G, Egger M, Pocock S J, Gøtzsche P C, Vandenbroucke J P, et al. Strengthening the reporting of observational studies in epidemiology (STROBE) statement: guidelines for reporting observational studies BMJ 2007; 335 :806 doi:10.1136/bmj.39335.541782.AD

